# Resting-State Amygdala Subregion and Precuneus Connectivity Provide Evidence for a Dimensional Approach to Studying Social Anxiety Disorder

**DOI:** 10.1101/2022.02.27.22271587

**Authors:** Simone Mizzi, Mangor Pedersen, Susan L Rossell, Peter Rendell, Gill Terrett, Markus Heinrichs, Izelle Labuschagne

## Abstract

**Background:** Social anxiety disorder (SAD) is a prevalent and disabling mental health condition, characterized by excessive fear and anxiety in social situations. Resting-state functional magnetic resonance imaging (fMRI) paradigms have been increasingly used to understand the neurobiological underpinnings of SAD in the absence of threat-related stimuli. Previous studies have primarily focused on the role of the amygdala in SAD. However, the amygdala consists of functionally and structurally distinct subregions, and recent studies have highlighted the importance of investigating the role of these subregions independently.

**Method:** Using multiband fMRI, we analyzed resting-state data from 135 participants (42 SAD, 93 healthy controls). By employing voxel-wise permutation testing, we examined group differences of fMRI connectivity and associations between fMRI connectivity and social anxiety symptoms to further investigate the classification of SAD as a categorical or dimensional construct.

**Results:** Seed-to-whole brain functional connectivity analysis using multiple ‘seeds’ including the amygdala and its subregions and the precuneus, revealed no statistically significant group differences. However, social anxiety severity was significantly negatively correlated with functional connectivity of the precuneus - perigenual anterior cingulate cortex and positively correlated with functional connectivity of the amygdala (specifically the superficial subregion) - parietal/cerebellar areas.

**Conclusion:** Our findings demonstrate clear links between symptomatology and brain connectivity in the absence of diagnostic differences, with evidence of amygdala subregion-specific alterations. The observed brain-symptom associations did not include disturbances in the brain’s fear circuitry (i.e., disturbances in connectivity between amygdala - prefrontal regions) likely due to the absence of threat-related stimuli.

## Introduction

Social anxiety disorder (SAD) is a debilitating mental health condition characterized by a disproportionate level of fear or anxiety in social situations that causes significant distress or functional impairment with a global lifetime estimated prevalence of 4.0% (1, 2). Accumulating evidence suggests that SAD may not exist as a discrete categorical entity (as defined by the Diagnostic and Statistical Manual of Mental Disorders, fifth edition; DSM-5) (1). Instead, it is proposed that symptoms associated with SAD have a dimensional structure (e.g., as a range of severity of anxiety, fear, and avoidance) (3-6). Given the high prevalence and subsequent impairments associated with SAD, there has been increased investigation to further understand the neurobiology of this disorder in the hope that it may improve its identification, classification, and treatment.

Advances in neuroimaging techniques have greatly assisted our understanding of the neurobiological mechanisms implicated in those with SAD. Most often, task-based functional magnetic resonance imaging (fMRI) approaches have elucidated the neural underpinnings of SAD under specific paradigms, such as in response to stimuli of facial expressions. In contrast, resting-state fMRI measures brain activity or connectivity in the absence of stimuli. In SAD, the use of resting-state fMRI allows for the identification of underlying neurobiological changes that are related to the characteristics of the disorder independent of any triggers from socially provoking situations (i.e., is the brain socially anxious outside the context of social threat?).

Findings from systematic reviews investigating task-based and resting-state fMRI in SAD have primarily identified the amygdala as a region of interest. That is, in response to socially relevant stimuli (i.e., threat-related facial expressions), those with SAD are commonly reported to have hyperactive amygdala responses compared to controls (7, 8). When examining only resting-state fMRI studies, we recently found that the most frequently reported alterations in connectivity were between amygdala-frontal regions in those with SAD compared to controls (9). However, there were mixed findings with regards to the direction of connectivity, with five studies reporting an increase (10-13) and four studies reporting a decrease (14-17).

There are several reasons to why these mixed findings may be occurring in the literature. Brain alterations at rest may depend on symptom severity, as supported by evidence of associations between social anxiety severity and resting-state functional connectivity of the amygdala-frontal regions in those with SAD (14, 18) and in a combined sample of participants with a diagnosis of SAD and those with no diagnosed psychiatric disorder (12). Additionally, mixed evidence regarding the connectivity patterns of the amygdala with frontal regions in those with SAD may relate to the amygdala having functionally distinct subregions (19-21). To date, two studies have examined resting-state amygdala subregion connectivity of the centromedial, superficial and basolateral complexes in SAD (11, 13). Both studies consistently found that those with SAD (compared to controls) had increased connectivity between each of the subregions and frontal regions (including the supplementary motor area, inferior frontal gyrus, superior frontal gyrus, dorsolateral prefrontal cortex, dorsomedial prefrontal cortex, and the anterior cingulate cortex). However, these findings and the majority of the previous studies showing that the amygdala and frontal regions are implicated in SAD have been limited in terms of interpretability for various reasons we discuss next.

Most studies to date have used relatively small sample sizes (average of n=23 SAD) which have been shown to detect unreliable brain and brain-behavior findings that are unlikely to be reproduced (22). Moreover, studies have used short resting-state scan lengths (ranging from 200-471 seconds) which have been associated with poor test-retest reliability of connectivity findings (23). Then, there has been considerable heterogeneity in scanning acquisition and pre-processing procedures which hinders the identification of consistencies in findings across the literature (9).

In the current study, we aimed to further elucidate resting-state fMRI connectivity differences in people with SAD compared to controls. We attempted to address the aforementioned limitations by including a larger sample size, a longer scan duration using multiband imaging which reduces the signal-to-noise ratio, and a streamlined and reliable data processing (fMRIPrep) pipeline (to allow for easier replication). Our specific aims were three-fold: i) to test whether resting-state functional connectivity from a range of ROIs or ‘seeds’ (including the amygdala and its subregions) displayed aberrant connectivity with other brain regions, in SAD compared to controls; ii) to examine a dimensional approach to the study of SAD by exploring the association between resting-state fMRI connectivity and social anxiety severity across all participants (SAD and controls), and iii) to test whether each subregion of the amygdala had functionally distinct connectivity patterns.

## Methods and Materials

### Participants

A total of 138 participants were included in this study, 43 of which had SAD and 95 of which were healthy controls. Participants were recruited using community-based advertising, and those with SAD were additionally recruited through online advertisements on the Anxiety Disorders Association of Victoria website and Facebook page.

Participants were included if they were aged between 18 to 55 years, fluent in English, and right-handed. They were excluded if they had a history of or current substance abuse (including smoking), head trauma (defined as being unconscious for ≥ 5 minutes), neurological condition, clinically significant medical illness (e.g., cardiovascular disease, diabetes), and MRI contraindications (e.g., metal objects that cannot be removed or unsafe for MRI. Additionally, the MINI 6.0.0 Screen (English version for the DSM-IV) was used to ensure that those in the control group had no prior or current psychiatric diagnosis. The MINI 6.0.0 English Version was used to determine whether those in the SAD group met the diagnostic criteria for SAD based on the DSM-IV or the DSM-5. The Liebowitz Social Anxiety Scale (LSAS) was used as an additional measure to determine whether participants in the clinical group met the diagnostic criteria for SAD, with a score ≥30 being required for inclusion (24, 25). The social interaction anxiety scale (SIAS) was administered to all participants as a measure of social anxiety symptom severity, with higher scores indicating greater levels of social anxiety (26). Written informed consent was obtained from all participants. This study was approved by the Human Research Ethics Committee at Australian Catholic University.

### Data Acquisition

Data acquisition was performed on a Siemens MAGNETOM Tim Trio 3.0 Tesla scanner with a Siemens 12 channel head matrix coil (Erlangen, Germany) at Swinburne University of Technology, Australia. Padded foam cushions were used to minimize head movement throughout the scan. All participants were instructed to try to think about nothing in particular (i.e., a resting-state), remain awake, and fixate their gaze on a white crosshair displayed centrally on a black background.

A multiband echo-planar imaging sequence with an acceleration factor of 5 was used to acquire functional MRI data for 8 minutes 38 seconds, along the anterior commissure-posterior commissure (AC-PC) plane with A > P phase encode direction (voxel size = 2 × 2 × 2 mm; 65 slices; repetition time (TR) = 1020 ms; total volumes = 500, echo time (TE) = 30 ms; flip angle (FA) = 65°). A T_1_-weighted sagittal MPRAGE structural image (TR = 1900 ms, TE = 2.52 ms, FA = 9°, 176 slices; voxel size = 1 × 1 × 1mm voxels) and T_2_-weighted image (TR = 3200 ms, TE = 402 ms, 176 slices; voxel size = 1 × 1 × 1 mm voxels) were also obtained for anatomical co-registration.

## Data Analysis

### Pre-Processing

T1- and T2-weighted MRI and resting-state fMRI images were converted to Brain Imaging Data Structure (BIDS) format (27). Firstly, the data was pre-processed using fMRIPrep 20.1.1 (Esteban, Markiewicz, et al. (2018); Esteban, Blair, et al. (2018); RRID:SCR_016216), based on Nipype 1.5.0 (Gorgolewski et al. (2011); Gorgolewski et al. (2018); RRID:SCR_002502) using the Ozstar High-Performance Computer (see Supplementary materials for details).

Following this, FSL was used to regress eight parameters out of the fMRI time series (signal from white matter, cerebrospinal fluid in addition to transverse x, y, and z head motion, and rotation x, y, and z head motion). Three participants (SAD = 1, Controls = 2) with excessive head motion, defined as a mean framewise displacement (FD) greater than 0.5 mm, were excluded from the study. This resulted in a total of 42 participants with SAD and 93 control participants. The data were filtered between 0.01 – 0.08 Hz and smoothed to 8 mm full width at half maximum (FWHM). Only fMRI voxels residing within grey matter were used for final analysis.

### Regions of Interest (ROIs)

Seed ROIs included the subregions of the amygdala. Additionally, regions that were frequently implicated in resting-state fMRI studies of SAD (as identified in the most recent systematic review on this topic (9)) were included to observe whether previous findings were replicable. ROIs were defined using two methods. The amygdala subregions were identified using cytoarchitectonic probability maps from the Anatomy Toolbox (Version 2.2b) in SPM12 (28, 29). All other ROIs (amygdala, subgenual anterior cingulate cortex (ACC), ventromedial prefrontal cortex (vmPFC), and the temporoparietal junction (TPJ)) were identified using NeuroSynth (http://neurosynth.org), which is an online database that uses a meta-analytic approach to synthesize existing neuroimaging literature (30). The relevant term was identified and the peak voxel of the region of interest was used as the MNI coordinate in this study (see Table 1).

**Table 1.** Coordinates and Size of Regions of Interests (ROIs)

| ROI |  | MNI<br>coordinates | Size of radius<br>sphere (mm) | How it was defined |
| --- | --- | --- | --- | --- |
| Amygdala: amygdalostriatal |  |  |  |  |
|  | Right | 27, -11, -11 | 3 | Anatomy toolbox |
|  | Left | -27, -11, -11 | 3 |  |
| Amygdala: basolateral |  |  |  |  |
|  | Right | 26, -5, -19 | 3 | Anatomy toolbox |
|  | Left | -26, -5, -19 | 3 |  |
| Amygdala: centromedial |  |  |  |  |
|  | Right | 23, -9, -10 | 3 | Anatomy toolbox |
|  | Left | -23, -9, -10 | 3 |  |
| Amygdala: superficial |  |  |  |  |
|  | Right | 19, -8, -14 | 3 | Anatomy toolbox |
|  | Left | -19, -8, -14 | 3 |  |
| Amygdala: whole amygdala |  |  |  |  |
|  | Right | 24, -4, -18 | 5 | Neurosynth: searched term<br>'emotional' <sup>b</sup> |
|  | Left | -24, -4, -18 | 5 |  |
| Precuneus |  |  |  |  |
|  | Right | 4, -58, 38 | 6 | Neurosynth: searched term<br>'default mode' <sup>a</sup> |
|  | Left | -4, -58, 38 | 6 |  |
| ACC (subgenual) |  |  |  |  |
|  | Right | 4, 32, -6 | 6 | Neurosynth: searched term<br>'emotional' <sup>b</sup> |
|  | Left | -4, 32, -6 | 6 |  |
| vmPFC |  |  |  |  |
|  | Right | 4, 48, -6 | 6 | Neurosynth: searched term<br>'default mode' <sup>a</sup> |
|  | Left | -4, 48, -6 | 6 |  |
| TPJ |  |  |  |  |
|  | Right | 56, -50, 16 | 6 | Neurosynth: searched term<br>'default mode' <sup>a</sup> |
|  | Left | -56, -50, 16 | 6 |  |

*Note.* <sup>a</sup> based on 777 studies and 26256 activations; <sup>b</sup> based on 1708 studies and 58327 activations ACC = anterior cingulate cortex; TPJ = temporoparietal junction; vmPFC = ventromedial prefrontal cortex.

### fMRI Connectivity Analysis

To assess whether group differences existed between the defined ROIs and the whole-brain, seed-based functional connectivity analysis was completed using Data Processing Assistant for Resting-State fMRI (DPARSF) V5.1 (31) (http://rfmri.org/DPARSF) within the Data Processing and Analysis for Brain Imaging (DPABI) (http://rfmri.org/DPAB) (32) implemented in MATLAB R2017b. Pre-processed voxel-wise fMRI time-series were extracted from each seed, and Pearson’s correlation coefficients were calculated between the average seed time series, and the time series of all voxels in the brain (total number of grey matter voxels = 173.843). The correlation coefficient was transformed to a *z-*value using Fisher’s r- to-z transformation, and the resultant functional connectivity maps for each participant was entered into the two-sample *t*-test.

## Statistical Analysis

To examine differences in age and mean FD between groups, the non-parametric Mann-Whitney *U* test was used as the data was not normally distributed (as determined by Shapiro-Wilk test; *p* < 0.001). To examine differences in SIAS scores and sex between groups, an independent sample *t*-test and a chi-square analysis were used respectively.

An independent sample *t*-test was used to analyze group differences in functional connectivity (SAD vs controls) for each ROI (aim i). Two-tailed threshold-free cluster enhancement (TFCE) correction with the Permutation Analysis of Linear Models (PALM) software (*p* < 0.05, 5000 permutations, tail approximation acceleration method) was applied (33, 34). Age, sex, and FD (average head movement) were included as covariates of no interest. Given the ongoing debate about the appropriateness of including covariates if they are not matched between groups (35), the *t*-tests were run a second time excluding FD as a covariate as head motion differed significantly between groups (control participants had greater head motion than SAD participants).

To examine associations between seeded fMRI connectivity maps and social anxiety severity (as measured by the SIAS) across all participants (aim ii), a voxel-wise Spearman’s partial correlation analysis was conducted (due to the non-normal distribution of social anxiety scores). Age and sex were included as covariates. Since TFCE does not support correlation-based permutations, we employed a *p-*min permutation approach to perform multiple comparisons testing (36, 37). In total, we randomized SIAS scores 5000 times (i.e., 5000 permutations) while keeping the fMRI connectivity data unchanged for all participants. For each permutation, we extract the minimal *p*-value across all 173,843 voxels, which in turn represents the null distribution (i.e., a ‘Bonferroni-like’ multiple comparison correction). This procedure asks the question: *what is the strongest correlation any voxel can have ‘by chance’*? The average of the 5000 random correlations obtained from the permutation testing was used to generate a statistical threshold. Any voxels with p-values less than this threshold were statistically significant. To avoid interpreting single voxels that may constitute a Type-1 error, we required 10 voxels to be interconnected to reach a statistical significance level. To ensure the functional specificity of amygdala subregions being measured (aim iii), average connectivity maps for each of the subregions (located in the left hemisphere) were compared to one another.

## Results

### Demographics

The demographic and clinical characteristics of all participants (aside from 3 participants who were excluded from all further analyses due to excessive head motion) are included in Table 2. Five participants in the SAD group had comorbid secondary psychiatric disorders (generalized anxiety disorder (n = 3); post-traumatic stress disorder (n = 1); obsessive-compulsive disorder/attention deficit hyperactivity disorder (n = 1)). There were no significant differences in age and sex between groups. There was a significant difference in mean FD (greater in-scanner head motion in the control group) and SIAS (higher scores in the SAD group). Of note, there was an overlap of SIAS scores between people who were diagnosed with SAD and controls (see the range in Table 2).

**Table 2.** Demographic Information of Participants

|  | SAD | Control | Statistic | *p*-value |
| --- | --- | --- | --- | --- |
| *n* | 42 | 93 | - | - |
| Sex<br>(female/male) | 21/21 | 49/44 | $\chi^2 = 0.084$ | 0.772 |
| Age<sup>a</sup> | 27.57 (7.52), 19.52 –<br>54.96 | 26.06 (6.50), 18.24 –<br>49.41 | $U =$<br>1718.000 | 0.264 |
| SIAS<sup>a</sup> | 51.62 (9.80), 28 – 72 | 18.29 (10.65), 1 – 48 | $t = 17.248$ | <0.001 |
| LSAS<sup>a</sup> | 81.24 (22.58), 41 – 139<sup>b</sup> | - | - | - |
| Mean FD<sup>a</sup> | 0.16 (0.07), 0.08 – 0.46 | 0.20 (0.08), 0.07 – 0.46 | $U =$<br>1289.000 | 0.002 |

*Note* . <sup>a</sup> Mean (standard deviation) and range; <sup>b</sup> scores were not reported for n=5; FD = framewise displacement; LSAS = Liebowitz Social Anxiety Scale; SIAS = Social Interaction Anxiety Scale; $t$ = independent sample $t$ -test; $\chi^2$ = chi square analysis; $U$ = Mann-Whitney $U$ test.

### No Between-Group Differences in Functional Connectivity

Seed-based functional connectivity from 18 ROIs showed no statistically significant group differences between SAD and controls, based on 5000 permutations. This was observed when average FD was included and excluded as a covariate of no interest. It is worth noting that we observed moderate, but sub-threshold, effect sizes between groups (see Supplementary Table 1 and Supplementary Figure 1).

### Significant Associations between Functional Connectivity and Social Anxiety Severity

In the combined groups, significant associations (*p* < 0.001, corrected for multiple comparisons using the *p*-min method) were found between SIAS scores and seeded fMRI connectivity of several amygdala subregions and the precuneus (see Table 3 and Figure 1).

**Table 3.** Significant Associations between Resting-State Functional Connectivity and Social Anxiety Severity (SIAS Scores)

| Seed region | Regions showing altered connectivity | Peak MNI coordinate | Peak intensity (Spearman $\rho$ value) | p-value | Correlation |
| --- | --- | --- | --- | --- | --- |
| L. superficial | R. supramarginal gyrus | 44 -46 36 | 0.361 | 0.00013 | Positive |
|  | L. cerebellum_crus2 | -8 -76 -30 | 0.360 | 0.00016 | Positive |
| L. amygdala | R. supramarginal gyrus | 44 -44 36 | 0.389 | 0.00016 | Positive |
|  | L. cerebellum_7b | -24 -70 -44 | 0.372 | 0.00014 | Positive |
| L. precuneus | R. peri-genu ACC (BA32) | 4 38 -10 | -0.396 | 0.00001 | Negative |
| R. precuneus | R. peri-genu ACC (BA32) | 4 38 -10 | -0.389 | 0.00013 | Negative |

*Note.* N = 135; ACC = anterior cingulate cortex; BA = Brodmann Area; L = left; MNI = Montreal Neurological Institute; SIAS = social interaction anxiety scale; R = right.

**Figure 1.**
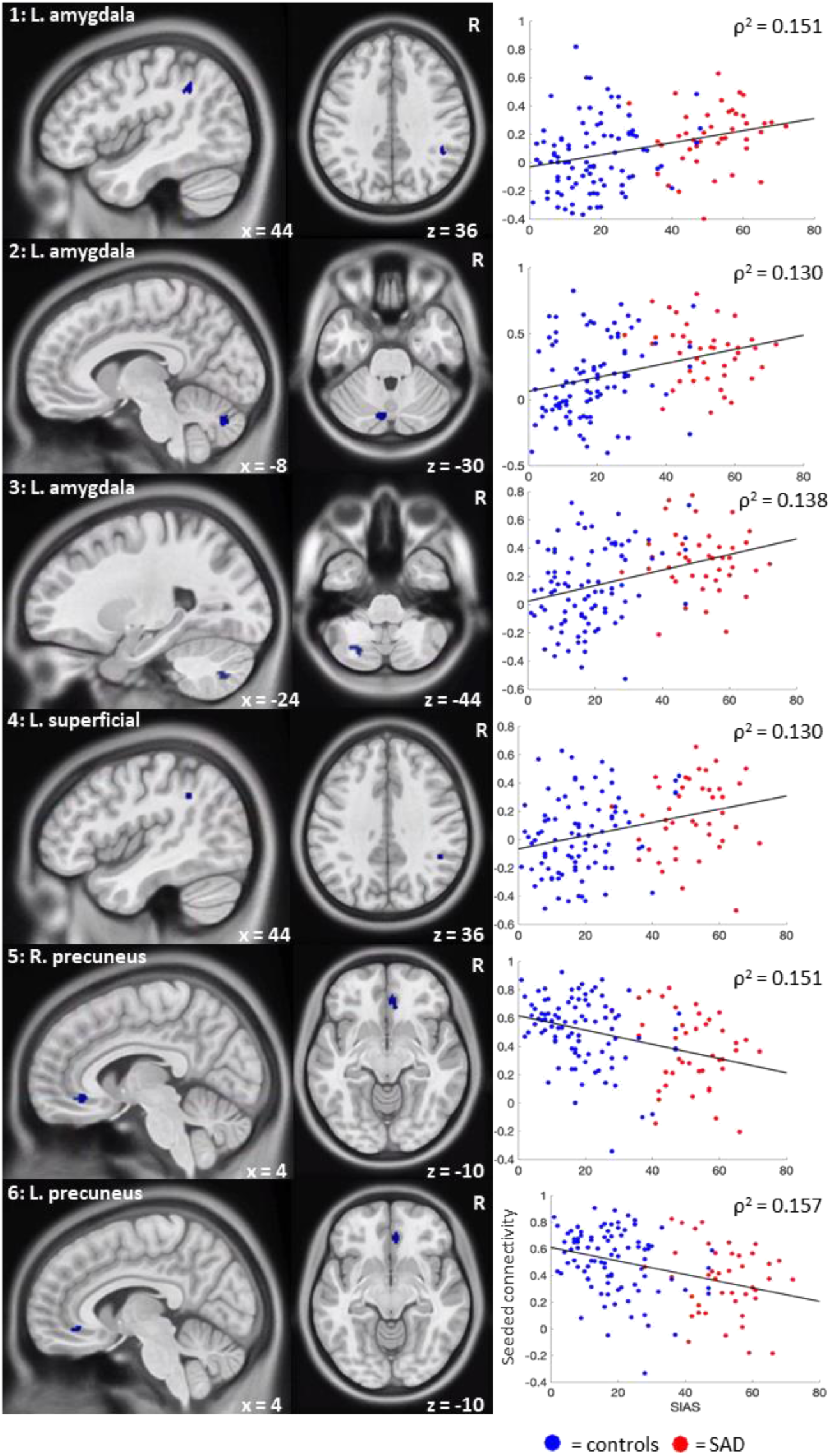
Connectivity maps and Spearman’s ρ correlations of the significant associations (all p < 0.001) between resting-state functional connectivity (y-axis) and social anxiety severity (SIAS scores; x-axis). Note. 1 = L. amygdala – R. supramarginal gyrus; 2 = L. amygdala – L. cerebellum_crus2; 3 = L. amygdala – L. cerebellum_7b; 4 = L. superficial – R. supramarginal gyrus; 5 = R. precuneus – R. peri-genu ACC ; 6 = L. precuneus – R. peri-genu ACC.

### Amygdala Subregions Display Divergent Functional Connectivity Patterns

In the combined groups, differences in the average connectivity maps when comparing amygdala subregions were observed and presented in Figure 2 (for illustration purposes we presented only the left hemispheric maps, but similar patterns were observed with right hemispheric subregions). These images show divergent connectivity patterns depending on the amygdala subregion ROI, thereby providing evidence that the connectivity maps from different amygdala subregions in our study are functionally distinct.

**Figure 2.**
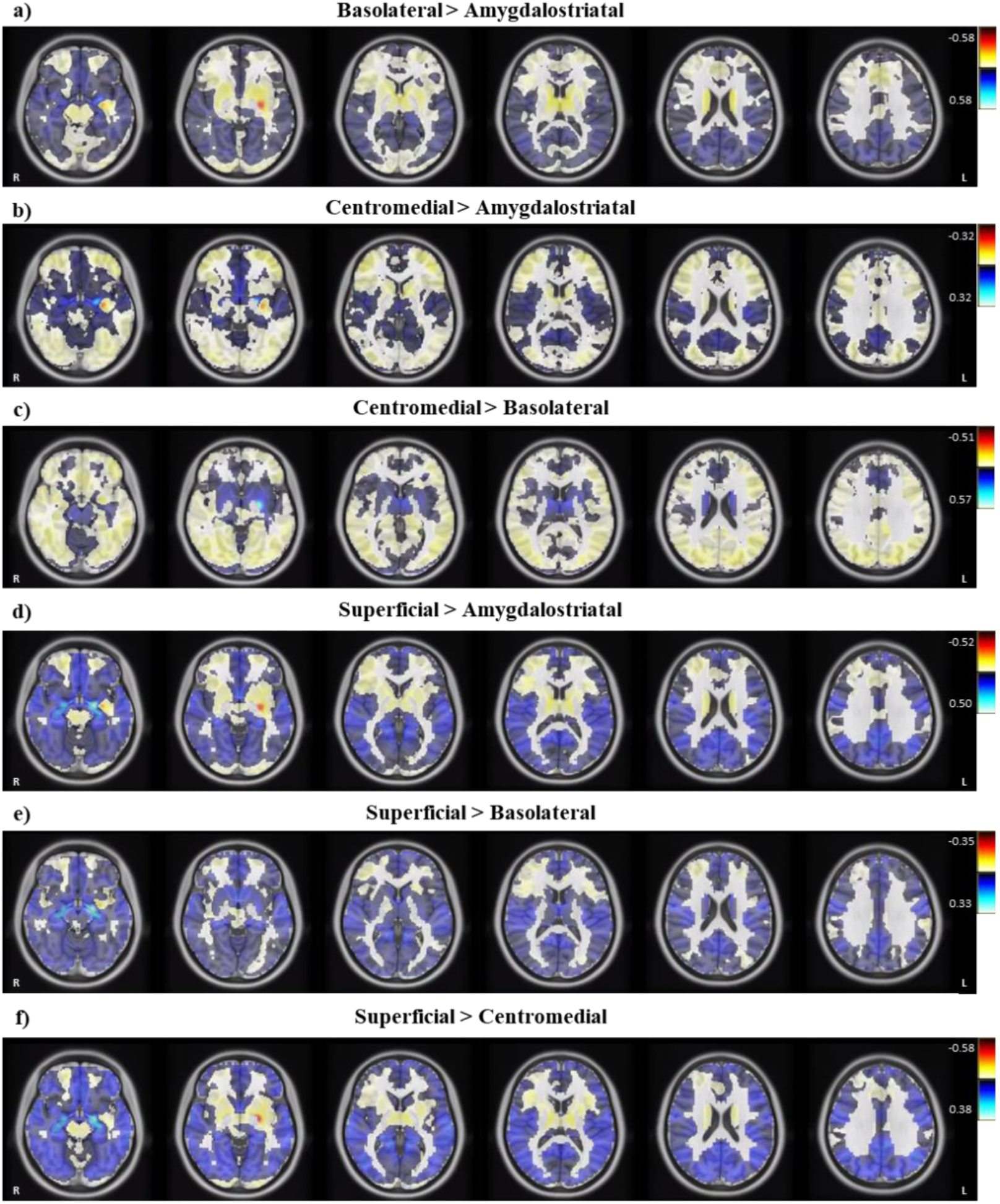
Differences in the Average Functional Connectivity Maps of Left Hemispheric Amygdala Subregions.

## Discussion

Using multiband fMRI, we found significant associations between social anxiety severity and resting-state functional connectivity across 135 participants (42 with SAD). Specifically, we found positive associations between severity of social anxiety and functional connectivity between the left superficial amygdala – right supramarginal gyrus and the left amygdala – right supramarginal gyrus and left cerebellar regions. We also found negative associations between social anxiety severity and resting-state functional connectivity of the bilateral precuneus and the right peri-genu ACC. These associations were observed in the absence of statistically significant group differences in resting-state functional connectivity (control vs. SAD participants).

### Amygdala subregion-specific associations with social anxiety

To date, very little is known about the functionality of the amygdala subregions in people with SAD given that no previous resting-state fMRI studies have examined these four commonly classified subregions within this population. Here, we showed that the positive association between social anxiety severity and functional connectivity between the left amygdala and the supramarginal gyrus is driven specifically by the superficial subregion of the amygdala. Increased connectivity between the superficial amygdala and the supramarginal gyrus at rest may indicate enhanced emotional surveillance of socially anxious self-relevant information and an increased tendency for socially anxious people to have negative social-evaluative cognitions, which is thought to maintain the disorder (i.e., negative thoughts/feelings they have about themselves are put onto others thus increasing feelings of fear/anxiety) (16, 38). This is because the superficial subregion of the amygdala has been implicated in the processing of socially relevant information (39). Additionally, the supramarginal gyrus is thought to play a role in downregulating egocentricity bias (i.e., the tendency to project one’s mental state onto others) (40), with evidence from a recent meta-analysis finding that those with SAD (compared to controls) had significantly decreased activation in this region when viewing disorder-related scenes (e.g., being in a conference room, harsh faces) compared to neutral scenes (41).

There were no significant associations between social anxiety severity and the amygdalostriatal, basolateral and centromedial subregions of the amygdala. This suggests that these subregions may play a role in socio-emotion processing (as measured by task-based fMRI studies) rather than in the absence of any stimuli (i.e., at rest). The centromedial subregion, which is known to be the major output area of the amygdala, sends signals to other neural areas to generate emotional, behavioral, autonomic, and motor responses (39). Therefore, aberrant patterns of connectivity in this subregion may only occur in task-based paradigms that require a response (and therefore an output signal). This is supported by findings of greater activation in the centromedial amygdala in response to negatively valenced stimuli compared to positively valenced stimuli in people genetically enriched for SAD (42) and significantly increased activation in the centromedial subregion of the amygdala in those with SAD (compared to controls) when viewing disorder-related scenes compared to neutral scenes (43). Similarly, the basolateral subregion is involved in the integration of sensory information from the environment (44). Less is known about the amygdalostriatal subregion, but it is thought to have shared pathways with the basolateral subregion (45). The absence of significant findings in these two subregions may reflect the lack of necessity to integrate information in the absence of stimuli during resting-state fMRI.

### Precuneus to ACC associations with social anxiety

In addition to the positive associations, we also observed negative associations between the connectivity of the precuneus to the peri-genu ACC and social anxiety severity. This finding is in line with Brühl, Delsignore (46) neurobiological model of SAD which posited a decrease in connectivity between the precuneus and ACC in those with SAD compared to controls. The precuneus plays a role in organizing attentional processes such as assessing the context of stimuli (47) and is a prominent component of the default mode network which is involved in self-referential processing (48). The peri-genu ACC is involved in emotion regulation and, in a recent meta-analysis, has been identified as having altered functional connectivity to other neural regions across anxiety and affective disorders (49). Disrupted connectivity between the peri-genu ACC and the precuneus is thought to be related to a decreased ability in being able to regulate negative self-relevant emotions which contributes to the maintenance of SAD, and has similarly been found in people with major depressive disorder (50).

### Theoretical implications

It is of note that, despite observing moderate effect sizes for group differences in functional connectivity, none of these findings remained significant after statistical thresholding *(p* < 0.05, 5000 permutations, TFCE corrected). Voxel-wise permutation testing, which was used in this study, has become an increasingly popular choice to deal with multiple comparisons that may occur in fMRI analyses. This is due to its high sensitivity and its recognition that voxels are not activated independently of their neighboring voxels (51, 52). However, it has also been found that using such a stringent thresholding approach has its limitations. In addition to fewer degree-of-freedom in TFCE between-group analysis compared to permutation-based correlation analysis, Noble, Scheinost (53) found that the TFCE approach was not able to detect any medium-sized effects in large sample sizes ranging from 480 to 493 healthy participants using the fMRI data in the Human Connectome Project. They concluded that numerous true effects may have been missed due to the prioritization of controlling family-wise error rates. The link between false-positive errors (which we can control) and false-negative errors (which we cannot control) is a non-trivial problem in contemporary science, but it remains imperative to minimize the former error type. Therefore, we believe that the differences reported in functional connectivity between groups in this study (but which did not survive thresholding) may be a relevant finding of interest and could be tested in a more hypothesis-driven way in future studies by pre-selecting voxels-of-interest or larger ROI based anxiety-specific a priori hypotheses which will result in fewer multiple comparisons.

Significant associations between functional connectivity and social anxiety severity (in the absence of significant group differences) further contribute to evidence of a dimensional or spectrum conceptualization of SAD, this time from a neurobiological perspective. This is consistent with the National Institute of Mental Health’s Research Domain Criteria (RDoC) framework which is advocating for a dimensional approach for the investigation of neurobiological markers of psychiatric disorders (54). Other studies that have used resting-state fMRI in a range of psychiatric and neurodevelopmental disorders (such as attention-deficit hyperactivity disorder, autism spectrum disorder, and major depressive disorder) have similarly found evidence to support the conceptualization of these disorders as being dimensional, with associations between functional connectivity and symptom severity (55-57). Findings from taxometric analyses (5) and previous fMRI studies in those with SAD also support this approach, with significant positive associations (but no significant group differences when comparing SAD to controls) between social anxiety severity and brain activity (e.g. in the dorsal ACC and right anterior insular cortex) in response to threat stimuli (58), and between emotion regulation and amygdala functional connectivity at rest (59).

Our findings of significant associations in the absence of statistically significant group differences (control vs. SAD participants) is also consistent with the most recently proposed integrated etiological and maintenance (IAM) model of SAD (38) in which contributing factors (both neurobiological and cognitive) to the etiology and maintenance of the disorder are identified as being dimensional. However, the most recently proposed neurobiological model of SAD (46) uses a categorical approach to conceptualize changes in neural activity and connectivity occurring in those with SAD compared to controls. Our findings show a similar pattern to the neurobiological model, including decreased connectivity between the precuneus and ACC in those with SAD compared to controls. However, our finding of increased connectivity between the amygdala (including the superficial subregion) and the supramarginal gyrus being associated with increased social anxiety severity is contrary to the model which indicates decreased connectivity between the amygdala and parietal regions in those with SAD compared to controls. Therefore, we provide further insights to this model by highlighting the importance of conceptualizing symptoms associated with SAD dimensionally (by examining associations and not only group comparisons) and the necessity to examine amygdala subregion specific effects that are linked to social anxiety severity. It is therefore critical that both these points are considered in future proposed neurobiological models of SAD.

It is well-known that broader disturbances between the amygdala and frontal regions are strongly implicated in fear processing, and altered connectivity between the amygdala and frontal regions in those with SAD compared to controls has been the most consistently reported across resting-state fMRI studies (reported by 9 of 18 fMRI studies in a systematic review) (9). However, we found no alterations in connectivity between amygdala-frontal regions between groups and no associations between amygdala-frontal connectivity and social anxiety severity. This suggests that people with social anxiety do not have disturbances in fear processing in the absence of explicit social stimuli (i.e., at rest), perhaps due to a lesser need to be hypervigilant to threat and a reduction in negative cognitions related to being evaluated by others (both factors contributing to the maintenance of social anxiety) (38).

## Conclusion

In conclusion, our study found significant associations between resting-state functional connectivity (with evidence of subregion-specific amygdala effects) and social anxiety severity scores in the absence of significant group differences. Relative to previous resting-state fMRI studies examining SAD, the strengths of this study were the use of a larger sample of participants (n=135) and longer scan length time (518 seconds; known to improve test-retest reliability) (60). Additionally, our use of multiband fMRI imaging (improving spatial and temporal resolution)(61), stringent fMRI thresholding, and use of *fMRIprep* for preprocessing provides a strong basis for future studies to continue studying and/or replicate these patterns. Based on the current findings, the IAM model of SAD (38), and the current RDoC framework, we believe that future studies would benefit from examining changes in brain activity and connectivity in relation to dimensional symptoms (e.g., social anxiety severity) rather than the presence or absence of a diagnosis of SAD (i.e., a categorical approach). This will lead to a more nuanced understanding of the neurobiological mechanisms underlying social anxiety at rest and may contribute to a future dimensional neurobiological model of SAD.

## Data Availability

All data produced in the present study are available upon reasonable request to the authors

## Acknowledgments and Disclosures

This work was supported by funding from the Australian Catholic University Research Fund Program Grant (ACURF2013000557), Australian Catholic University Early Career Research Fund (14HS4027IL), and by the Australian Government Research Training Program Scholarship (S.M.). S.L.R holds a Senior National Health and Medical Research Council (NHMRC) Fellowship (GNT1154651). The authors report no biomedical financial interests or potential conflicts of interest.

