## Supplementary Information for "Resting-State Amygdala Subregion and Precuneus Connectivity Provide Evidence for a Dimensional Approach to Studying Social Anxiety Disorder"

### Methods

#### fMRIprep Pre-Processing

##### *Anatomical Data reprocessing*

T1-weighted (T1w) images were corrected for intensity non-uniformity (INU) with N4BiasFieldCorrection (1), distributed with ANTs 2.2.0 (2) (RRID:SCR_004757), and used as T1w-reference throughout the workflow. The T1w-reference was then skull-stripped with a Nipype implementation of the antsBrainExtraction.sh workflow (from ANTs), using OASIS30ANTs as target template. Brain tissue segmentation of cerebrospinal fluid (CSF), white-matter (WM) and gray-matter (GM) was performed on the brain-extracted T1w using fast (FSL 5.0.9, RRID:SCR_002823) (3). Brain surfaces were reconstructed using recon-all (FreeSurfer 6.0.1, RRID:SCR_001847)(4), and the brain mask estimated previously was refined with a custom variation of the method to reconcile ANTs-derived and FreeSurfer-derived segmentations of the cortical gray-matter of Mindboggle (RRID:SCR_002438)(5).

Volume-based spatial normalization to two standard spaces (MNI152NLin6Asym, MNI152NLin2009cAsym) was performed through nonlinear registration with antsRegistration (ANTs 2.2.0), using brain-extracted versions of both T1w reference and the T1w template. The following templates were selected for spatial normalization: FSL's MNI ICBM 152 non-linear 6th Generation Asymmetric Average Brain Stereotaxic Registration Model [(6), RRID:SCR_002823; TemplateFlow ID: MNI152NLin6Asym], ICBM 152 Nonlinear Asymmetrical template version 2009c [(7), RRID:SCR_008796; TemplateFlow ID: MNI152NLin2009cAsym],

##### *Functional Data Preprocessing*

For each of the BOLD runs per subject (across all tasks and sessions), the following preprocessing was performed. First, a reference volume and its skull-stripped version were generated using a custom methodology of fMRIPrep. Head-motion parameters with respect to the BOLD reference (transformation matrices, and six corresponding rotation and translation parameters) are estimated before any spatiotemporal filtering using mcflirt (FSL 5.0.9)(8). BOLD runs were slice-time corrected using 3dTshift from AFNI 20160207 (9) (RRID:SCR_005927).

A deformation field to correct for susceptibility distortions was estimated based on fMRIPrep's fieldmap-less approach. The deformation field is that resulting from co-registering the BOLD reference to the same-subject T1w-reference with its intensity inverted (10, 11). Registration is performed with antsRegistration (ANTs 2.2.0), and the process regularized by constraining deformation to be nonzero only along the phase-encoding direction, and modulated with an average fieldmap template (12). Based on the estimated susceptibility distortion, a corrected EPI (echo-planar imaging) reference was calculated for a more accurate co-registration with the anatomical reference. The BOLD reference was then co-registered to the T1w reference using bbregister (FreeSurfer) which implements boundary-based registration (13).

Co-registration was configured with six degrees of freedom. The BOLD time-series were resampled onto the following surfaces (FreeSurfer reconstruction nomenclature): fsaverage6. The BOLD time-series (including slice-timing correction when applied) were resampled onto their original, native space by applying a single, composite transform to correct for head-motion and susceptibility distortions. These resampled BOLD time-series will be referred to as preprocessed BOLD in original space, or just preprocessed BOLD. The BOLD time-series were resampled into standard space, generating a preprocessed BOLD run in MNI152NLin6Asym space.

Several confounding time-series were calculated based on the preprocessed BOLD: framewise displacement (FD), DVARS and three region-wise global signals. FD was computed using two formulations following Power (absolute sum of relative motions, (14)) and Jenkinson (relative root mean square displacement between affines, (8)). FD and DVARS are calculated for each functional run, both using their implementations in Nipype (following the definitions by (14)). The three global signals are extracted within the CSF, the WM, and the whole-brain masks. Additionally, a set of physiological regressors were extracted to allow for component-based noise correction (CompCor, (15)). Principal components are estimated after high-pass filtering the preprocessed BOLD time-series (using a discrete cosine filter with 128s cut-off) for the two CompCor variants: temporal (tCompCor) and anatomical (aCompCor). tCompCor components are then calculated from the top 5% variable voxels within a mask covering the subcortical regions. This subcortical mask is obtained by heavily eroding the brain mask, which ensures it does not include cortical GM regions. For aCompCor, components are calculated within the intersection of the aforementioned mask and the union of CSF and WM masks calculated in T1w space, after their projection to the native space of each functional run (using the inverse BOLD-to-T1w transformation). Components are also calculated separately within the WM and CSF masks. For each CompCor decomposition, the k components with the largest singular values are retained, such that the retained components' time series are sufficient to explain 50 percent of variance across the nuisance mask (CSF, WM, combined, or temporal). The remaining components are dropped from consideration.

The head-motion estimates calculated in the correction step were also placed within the corresponding confounds file. The confound time series derived from head motion estimates and global signals were expanded with the inclusion of temporal derivatives and quadratic terms for each (16). Frames that exceeded a threshold of 0.5 mm FD or 1.5 standardised DVARS were annotated as motion outliers. All resamplings can be performed with a single interpolation step by composing all the pertinent transformations (i.e. head-motion transform matrices, susceptibility distortion correction when available, and co-registrations to anatomical and output spaces). Gridded (volumetric) resamplings were performed using antsApplyTransforms (ANTs), configured with Lanczos interpolation to minimize the smoothing effects of other kernels (17). Non-gridded (surface) resamplings were performed using mri_vol2surf (FreeSurfer).

### Results

**Table S1**

*Group Differences in Resting-State Functional Connectivity between those with SAD (n = 42) and Controls (n = 93)*

| Seed # | Seed region | SAD vs. Controls | | |
|---|---|---|---|---|
| | | **Regions showing peak altered connectivity** | **Peak MNI coordinate** | **Peak intensity** |
| 1 | L. amygdalostriatal | R. calcarine gyrus | 20 -90 4 | 3.892 |
| 2 | R. amygdalostriatal | R. superior frontal gyrus | 18 56 12 | 3.960 |
| 3 | L. basolateral | R. supramarginal gyrus (inferior parietal lobule) | 52 -38 42 | 4.1655 |
| 4 | R. basolateral | L. cerebellum (IV-V) | -18 -28 -26 | 4.5144 |
| 5 | L. centromedial | R. medial temporal pole | 46 16 -42 | 3.7061 |
| 6 | R. centromedial | R. superior frontal gyrus | 18 56 12 | 3.6414 |
| 7 | L. superficial | R. supramarginal gyrus (inferior parietal lobule) | 58 -38 34 | 4.0305 |
| 8 | R. superficial | L. anterior agranular insula complex | -30 -62 0 | 3.4051 |
| 9 | L. amygdala | R. supramarginal gyrus (inferior parietal lobule) | 52 -40 42 | 4.1854 |
| 10 | R. amygdala | L. cerebellum (IV-V) | -20 -28 -26 | 3.7871 |
| 11 | L. precuneus | R. calcarine gyrus | 22 -90 2 | 4.0954 |
| 12 | R. precuneus | R. frontal opercular area 2 | 20 -60 42 | 3.9529 |
| 13 | L. ACC (subgenual) | R. cerebellum (IX) | 2 -50 -42 | -4.1908 |
| 14 | R. ACC (subgenual) | R. cerebellum (IX) | 4 -48 -42 | -3.7414 |
| 15 | L. vmPFC | L. cingulate gyrus, frontal opercular area 1 | -16 8 46 | -4.294 |
| 16 | R. vmPFC | L. cingulate gyrus, frontal opercular area 1 | -16 8 46 | -3.8349 |
| 17 | L. TPJ | R. anterior agranular insula complex | -42 -26 0 | -4.3152 |
| 18 | R. TPJ | L. precuneus, frontal opercular area 3 | -18 -52 56 | 3.8643 |

*Note.* Region names were identified using the Automatic Anatomical Labelling Atlas and Glasser, Coalson (18) parcellation map. L = left; R = right; MNI = Montreal Neurological Institute; ACC = anterior cingulate gyrus; PFC = prefrontal cortex; TPJ = temperoparietal junction

**Figure S1**. *Seed-based functional connectivity maps of 18 seed regions of 42 SAD compared to 93 control participants.*

**1. Left amygdalostriatal**

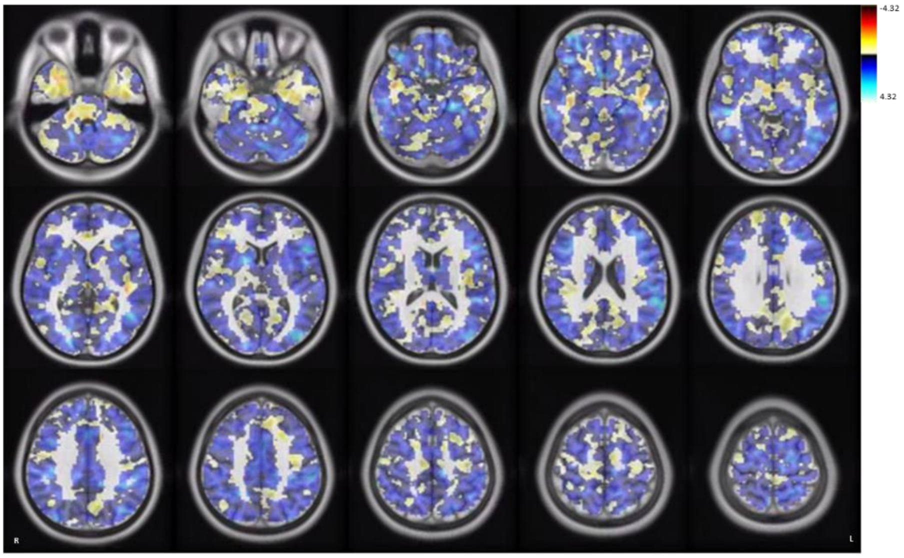


**2. Right amygdalostriatal**

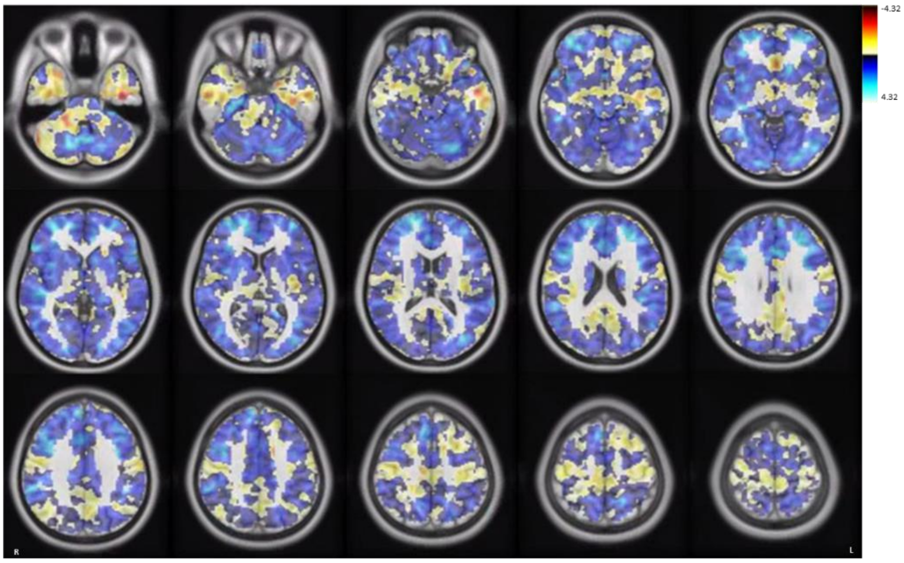


**3. Left basolateral**

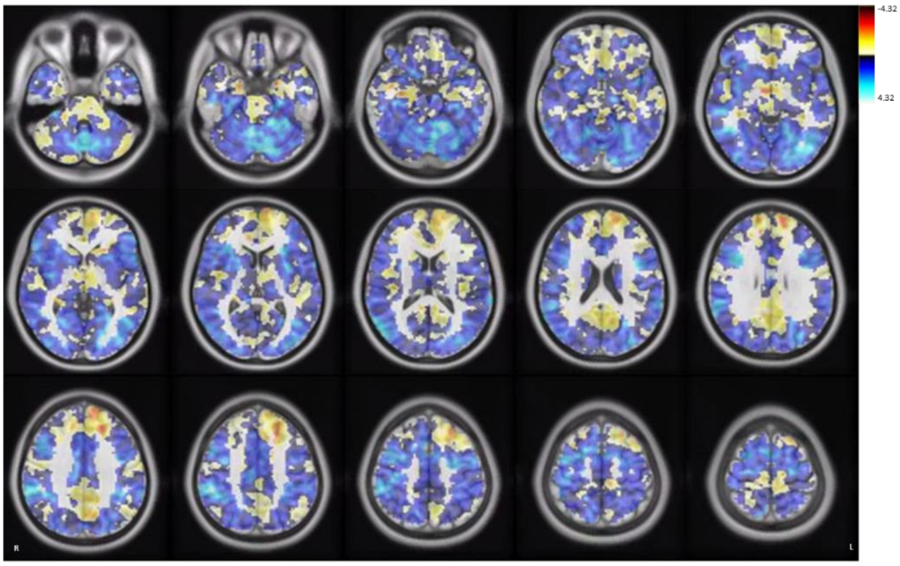

**4. Right basolateral**

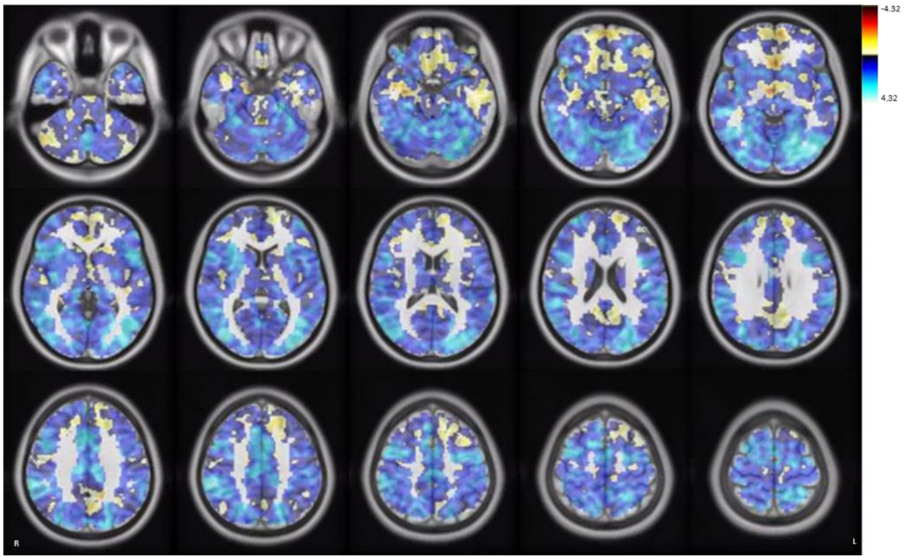


**5. Left centromedial**

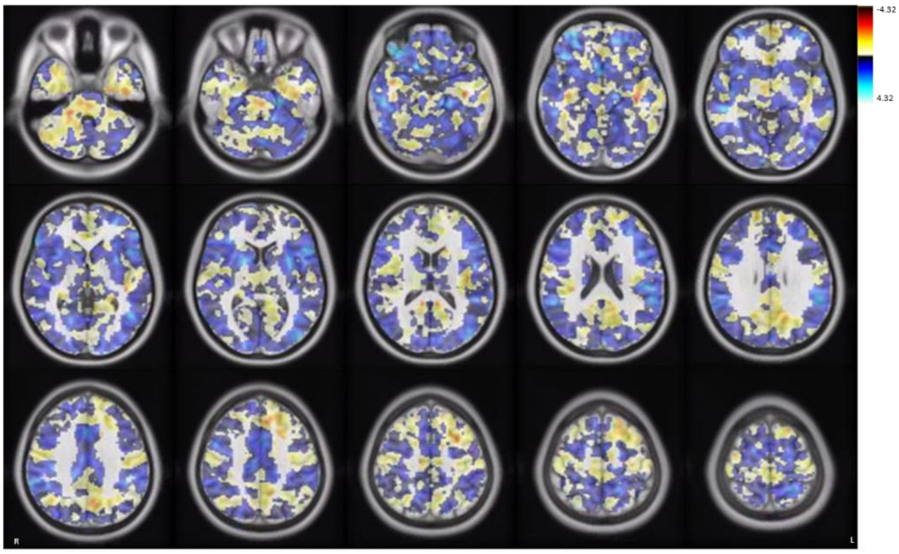


**6. Right centromedial**

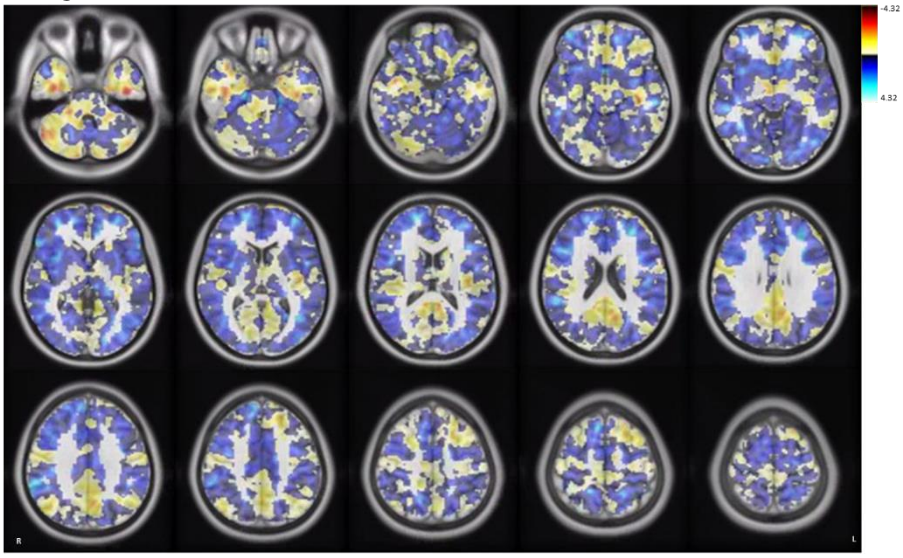

**7. Left superficial**

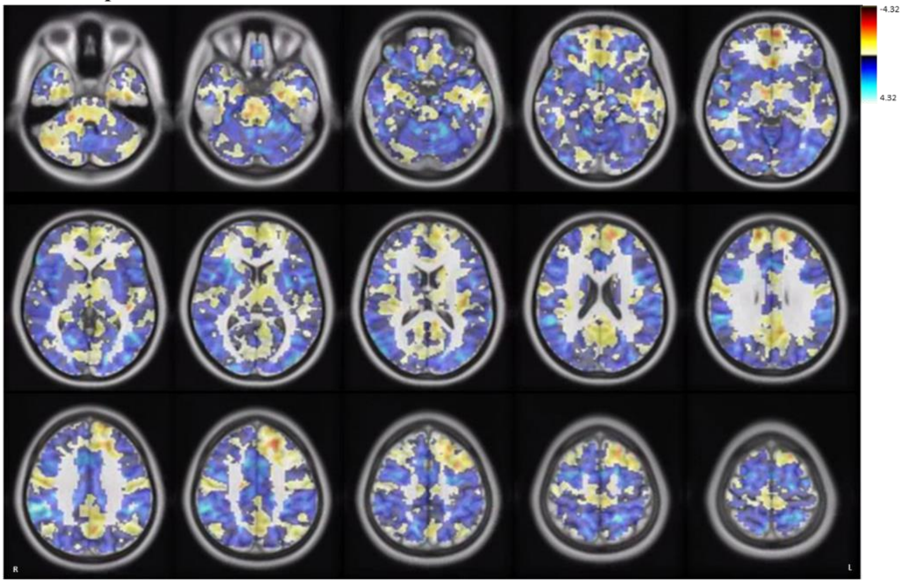


**8. Right superficial**

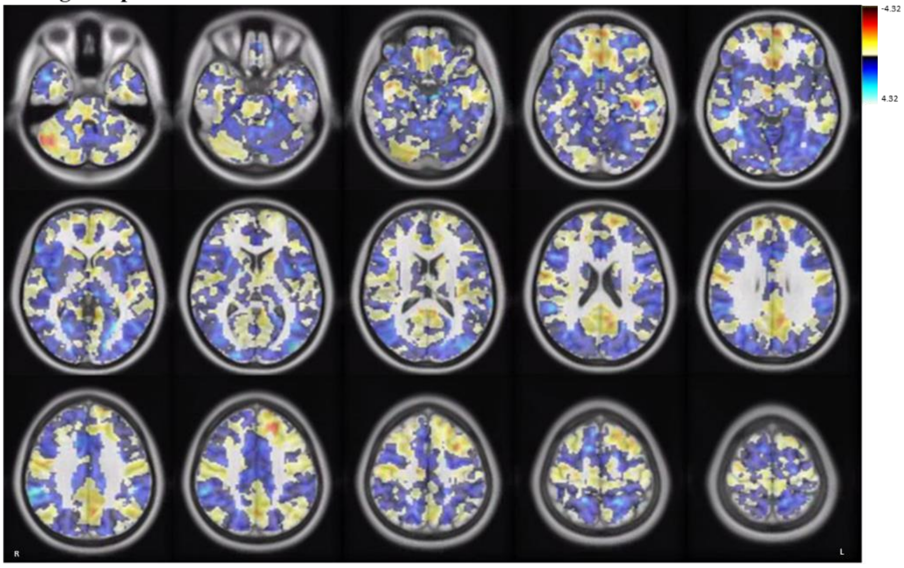


**9. Left amygdala**

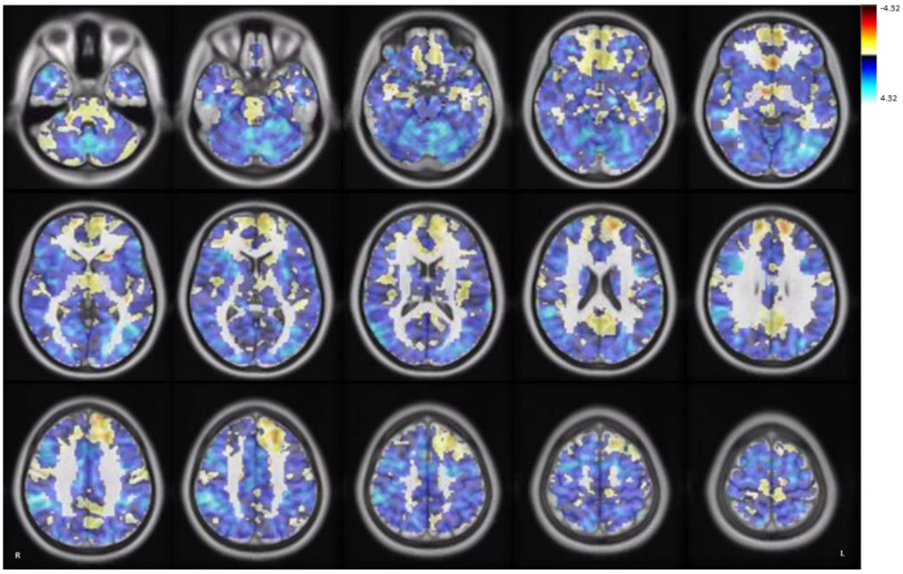

**10. Right amygdala**

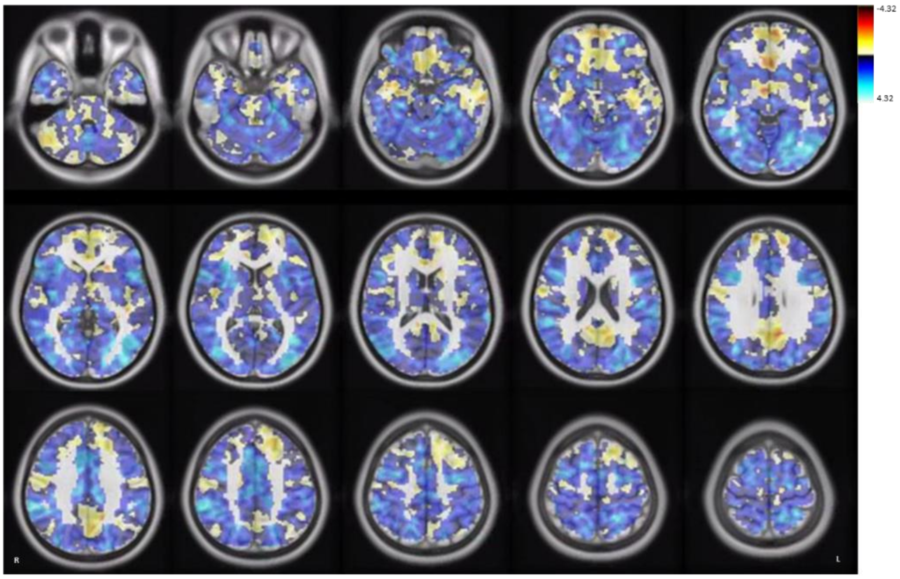


**11. Left precuneus**

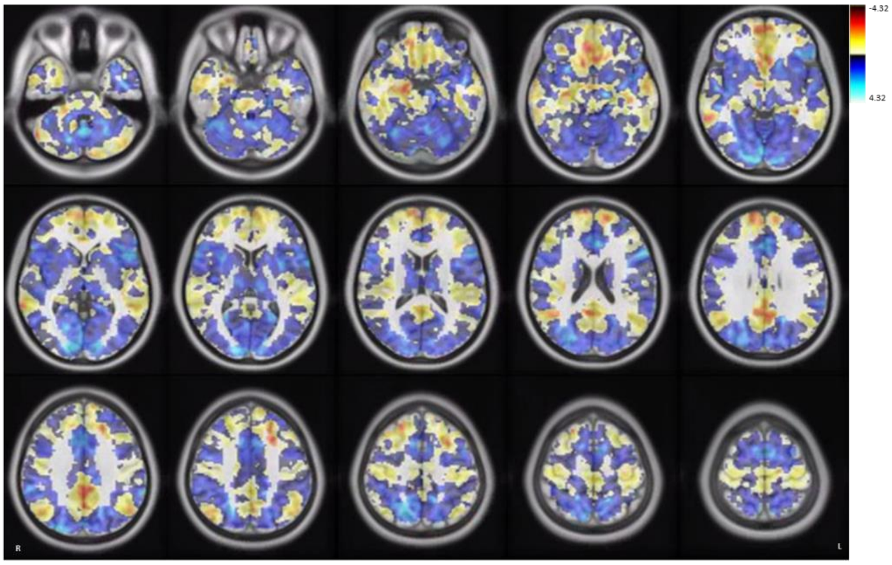


**12. Right precuneus**

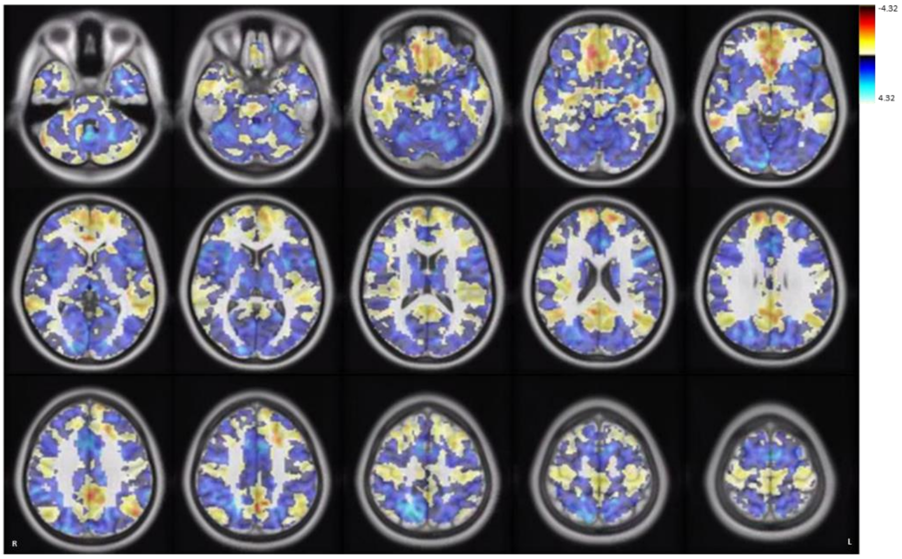

**13. Left anterior cingulate cortex (subgenual)**

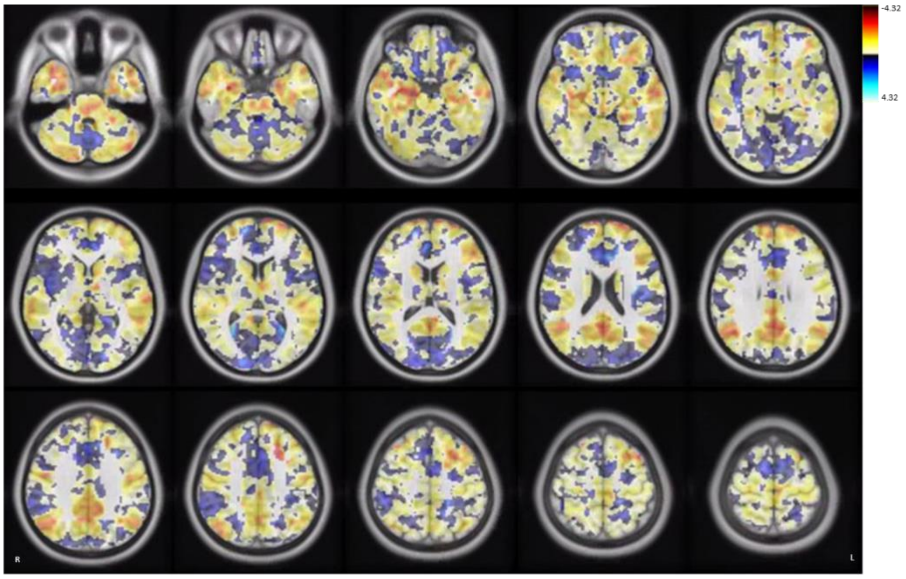


**14. Right anterior cingulate cortex (subgenual)**

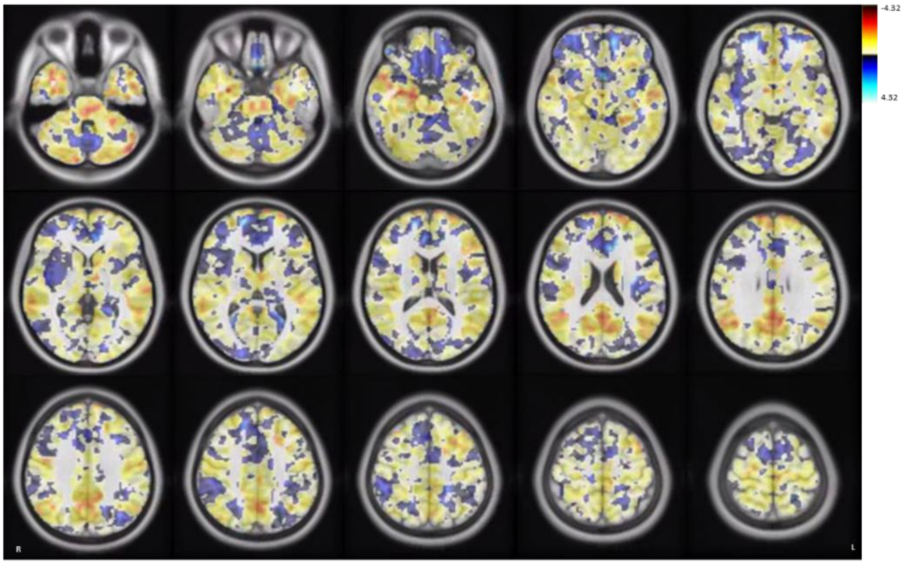


**15. Left ventromedial prefrontal cortex**

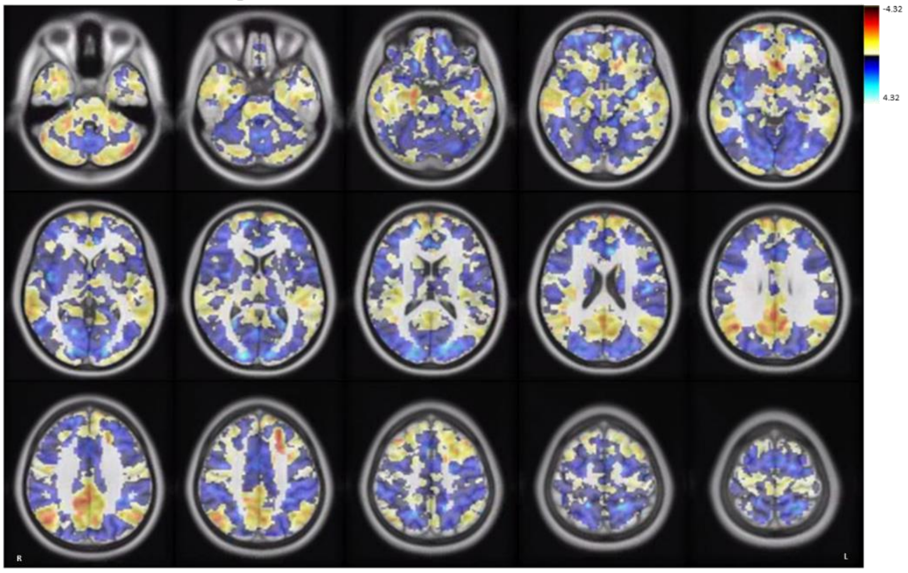

**16. Right ventromedial prefrontal cortex**

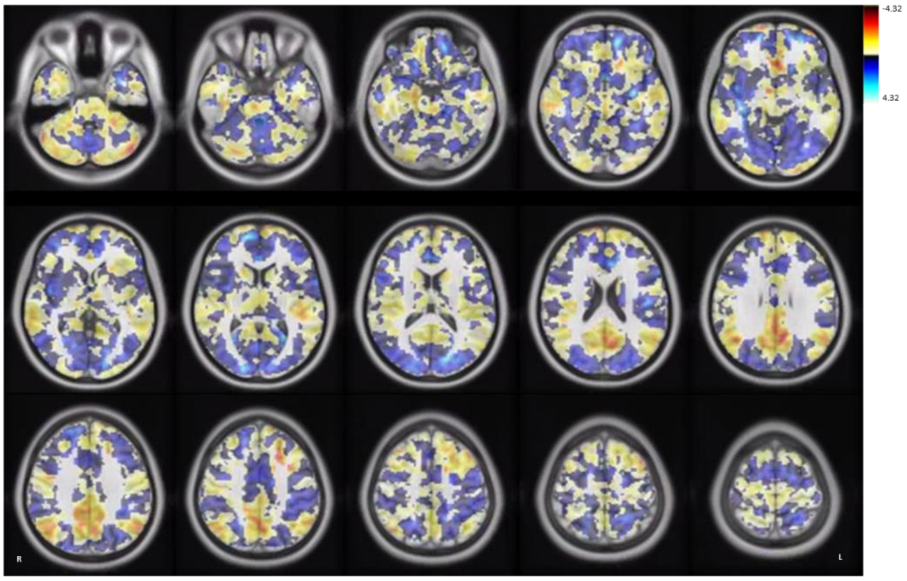


**17. Left temporoparietal junction**

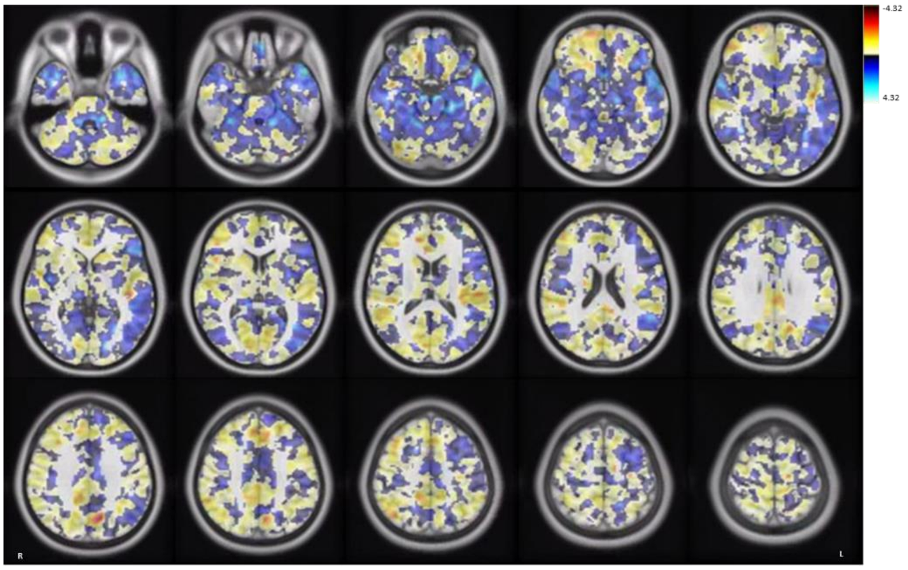


**18. Right temporoparietal junction**

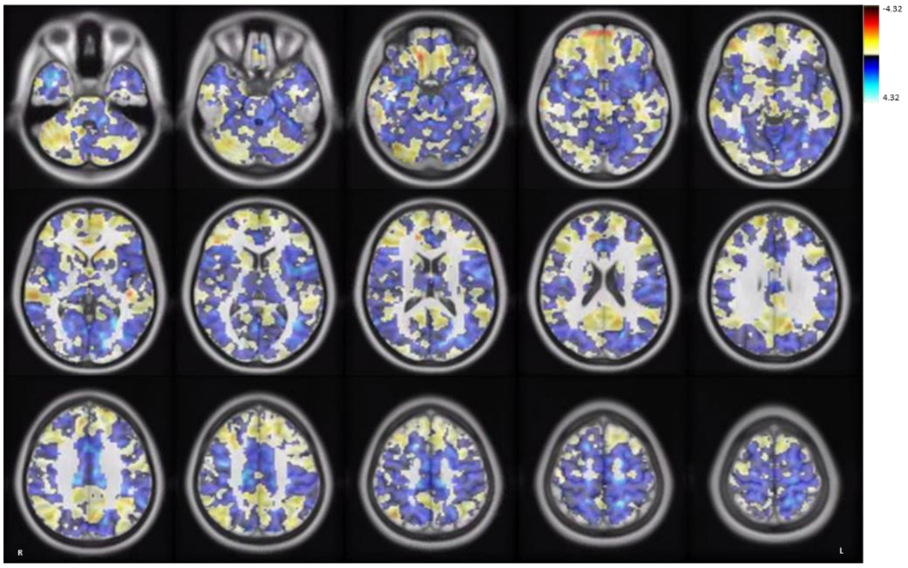
